# Computation of Socio-Economic Status in the AWI-Gen Project

**DOI:** 10.1101/2024.08.22.24312411

**Authors:** Scott Hazelhurst, Palwendé Boua, Ananyo Choudhury, Siyanda Madala, Dhriti Sengupta, Furahini Tluway, Michèle Ramsay

## Abstract

Socio-economic status of participants in many public health, epidemiological, and genome-wide association studies is an important trait of interest. It is often used in these studies as a measure of direct interest or as a covariate. The Africa Wits INDEPTH Partnership for Genomic and Environmental Research (AWI-Gen) explores genomic and environmental factors in non-communicable diseases, particularly cardio-metabolic disease. In Phase I of AWI-Gen, approximately 12,000 participants were recruited at six sites in four African countries. Participants were asked questions about asset ownership. This technical note describes how AWI-Gen computed socio-economic status from the asset register.

## 1. Introduction

Computing socio-economic status (SES) in many low- and middle-income countries is difficult because of lack of data, particularly around income and consumption. A common approach taken by health and demographic surveillance sites (HDSSs) is to use asset registers as a proxy: a suitable register of assets is chosen, and participants are asked to indicate yes/no whether they own the asset. From these data, we can compute an SES score for each individual. Commonly, for many analyses the participants are ranked into quintiles. If a study is cross-site, analyses are typically done per site — since assets mean different things in different communities, comparing absolute between sites is difficult. Using this method, we have the relative wealth of the person to the rest of the community as our measure.

The question addressed in this note is how to compute the per-individual SES score from the asset register.

- The obvious (and simplistic approach), which we call the *raw score*, is to code *yes*=1 and *no*=0 and to sum the assets that an individual owns.
- An alternative approach favoured by many HDSSs involves a principal component analysis of the SES variables (household assets), predicting factor scores, and recomputing a score using these as weights (described in more detail below).

The PCA method has become popular in HDSS work, popularised and extended by work such as [1, 2, 3]. The goal of the technique is to combine the asset register values in a sensible way. One of the issues with the asset register is that there is correlation between the items, and so just adding values may lead to a distorted view. By comparing the relationship of the asset register variables with each other through a PCA we can capture the independent parts of the assets.

The AWI-Gen Collaborative Centre [4] has used an asset register to estimate SES. Despite the attractiveness of the PCA method, we used the raw score. In a previous paper on methods [5], we incorrectly stated that we had used the PCA method. We have submitted a correction to the journal concerned and document what we did in this note.

## 2. Technical definitions

### 2.1. Asset register

We start with the asset register, with *m* individuals (rows) and *n* assets (columns).

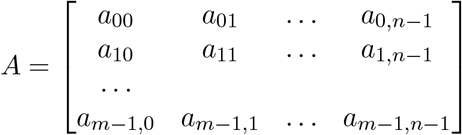

*a*_*ij*_ ∈ {0, 1} indicates whether person *i* has asset *j*. Since we do all analyses per site, we assume all individuals come from one site, and that the asset register is applicable to all individuals.

### 2.2. Raw score

We formally define the raw score of an indidual as

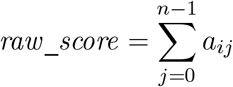

### 2.3. Principal component analysis

The goal of the PCA is to understand the relationship of the assets with each to find correlations between them. This done by:

- Computing the correlation matrix of the assets, or alternatively normalising by asset and computing the covariance matrix of the transpose of the matrix. (To use the covariance matrix, compute *µ_j_* as the mean of the asset *j* (column *j*), *σ*_*j*_ as the standard deviation of the asset *j* (column *j*) and then define *Â* by 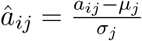.)
- Let **C** = Cov(*Â*^⊺^) be the covariance matrix of the transpose of the normalised asset register. Note that this is an *n* × *n* matrix since it captures the relationship of assets to each other.
- In order to manage the correlation of the different assets we now perform principal component analysis. Let **ev**_1_, **ev**_2_, … **ev**_*n*_ be the eigenvectors of **C**, and λ_1_, …, λ_*n*−1_ be the corresponding eigenvalues, ordered by descending size of eigenvalue.
- We follow [2] and use only the first eigenvector: for this reason the first eigenvalue is only used to indicate the percentage variation captured – methods that use multiple PCs will use the eigenvalues in this score computation.
- The first eigenvector **ev**_1_ = ⟨*w*_0_, … *w*_*n*−1_⟩ represents the weights of the different assets according to a PC analysis. The individual *w*_*i*_ are called variable *loadings* – the impact of variable on the first PC (also known as *factor*).

### 2.4. PCA adjusted score

We use the first eigenvector to compute the PCA adjusted score:

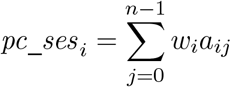

As mentioned, this can be generalised to using multiple eigenvectors (e.g. [3]) but it appears more common to use only one PC.

### 2.5. Compute the quintiles

We compute the quintiles from both the raw and adjusted scores.

## 3. Comparison on AWI-Gen Phase 1 data

We computed raw and PCA-based SES scores and grouped individuals by quintile. For the PCA scores we used the method described in Section 2.3.

### 3.1. Correlations

The correlations between the raw and PCA-SES scores are shown in the table below per site. The column *SES-Cor* shows the Pearson correlation between the raw SES score and the PC SES score. The column *SES-Q-R* shows the Spearman correlation between the corresponding quintiles computed with the two methods.

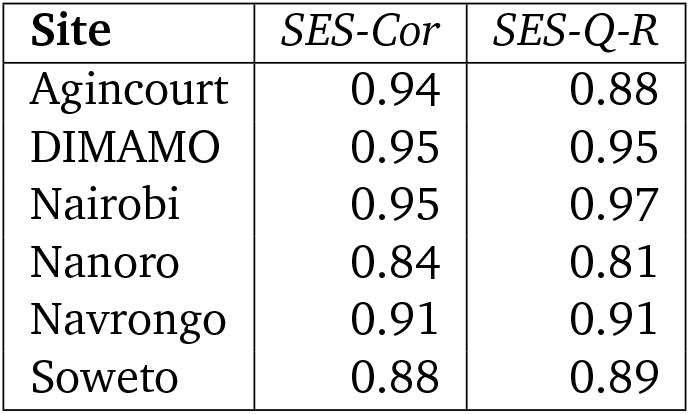

| Site | *SES-Cor* | *SES-Q-R* |
| --- | --- | --- |
| Agincourt | 0.94 | 0.88 |
| DIMAMO | 0.95 | 0.95 |
| Nairobi | 0.95 | 0.97 |
| Nanoro | 0.84 | 0.81 |
| Navrongo | 0.91 | 0.91 |
| Soweto | 0.88 | 0.89 |

Figures 1–6 visually show the corresponding raw and PC estimations of SES using the first PC. The factor loadings (i.e., the indivdiual elements of the eigenvector) for the first principal component were computed and are shown in each figure too.

**Figure 1:**
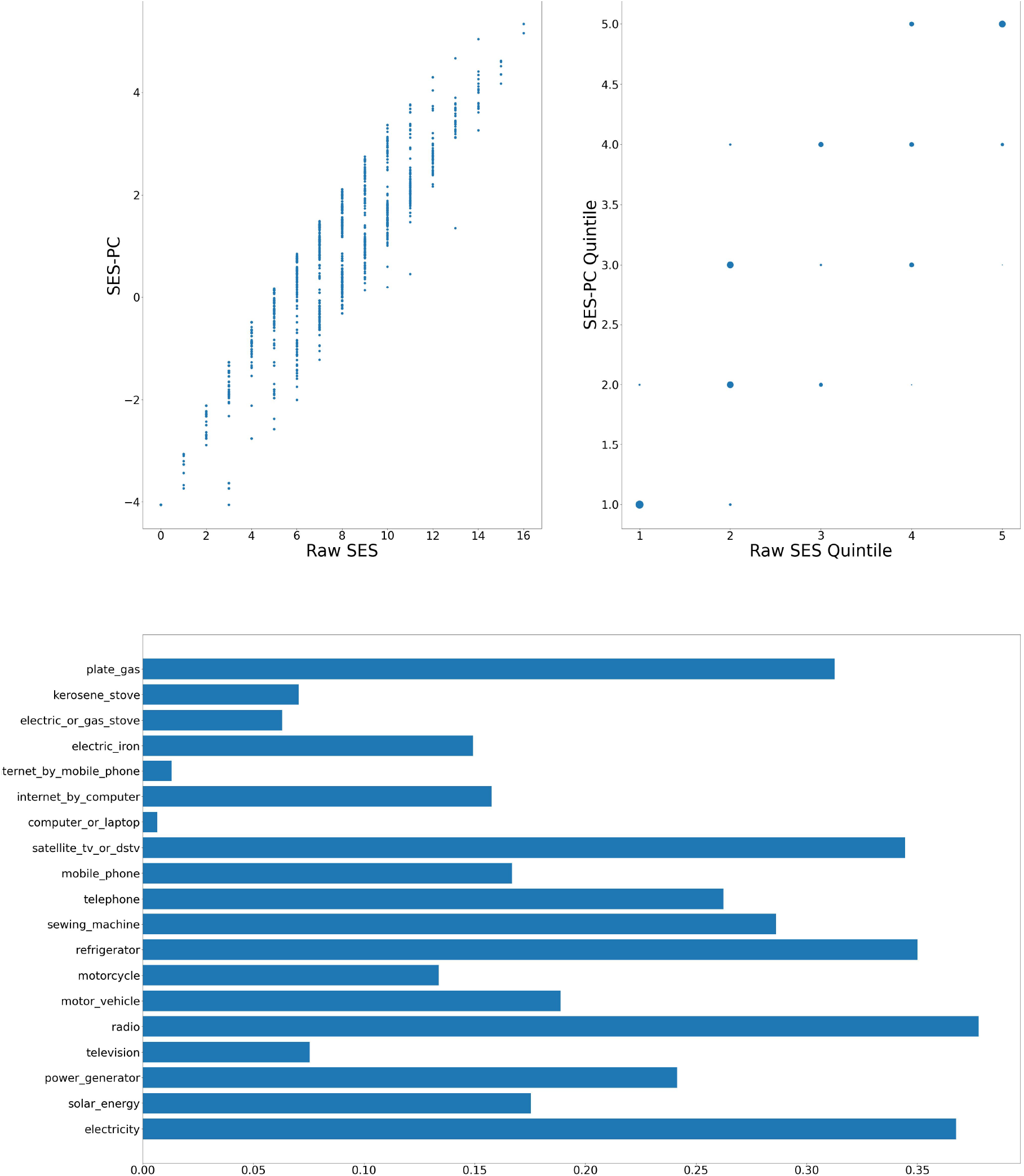
Agincourt — Computation of SES score: graph at top left shows a scatter plot of the SES raw value versus adjusted value; top right shows the scatter plot of the computation quintiles using the different method with the size of the dot reflecting the number of individuals in each category; and at the bottom, the resulting weights of the assets for Agincourt.

**Figure 2:**
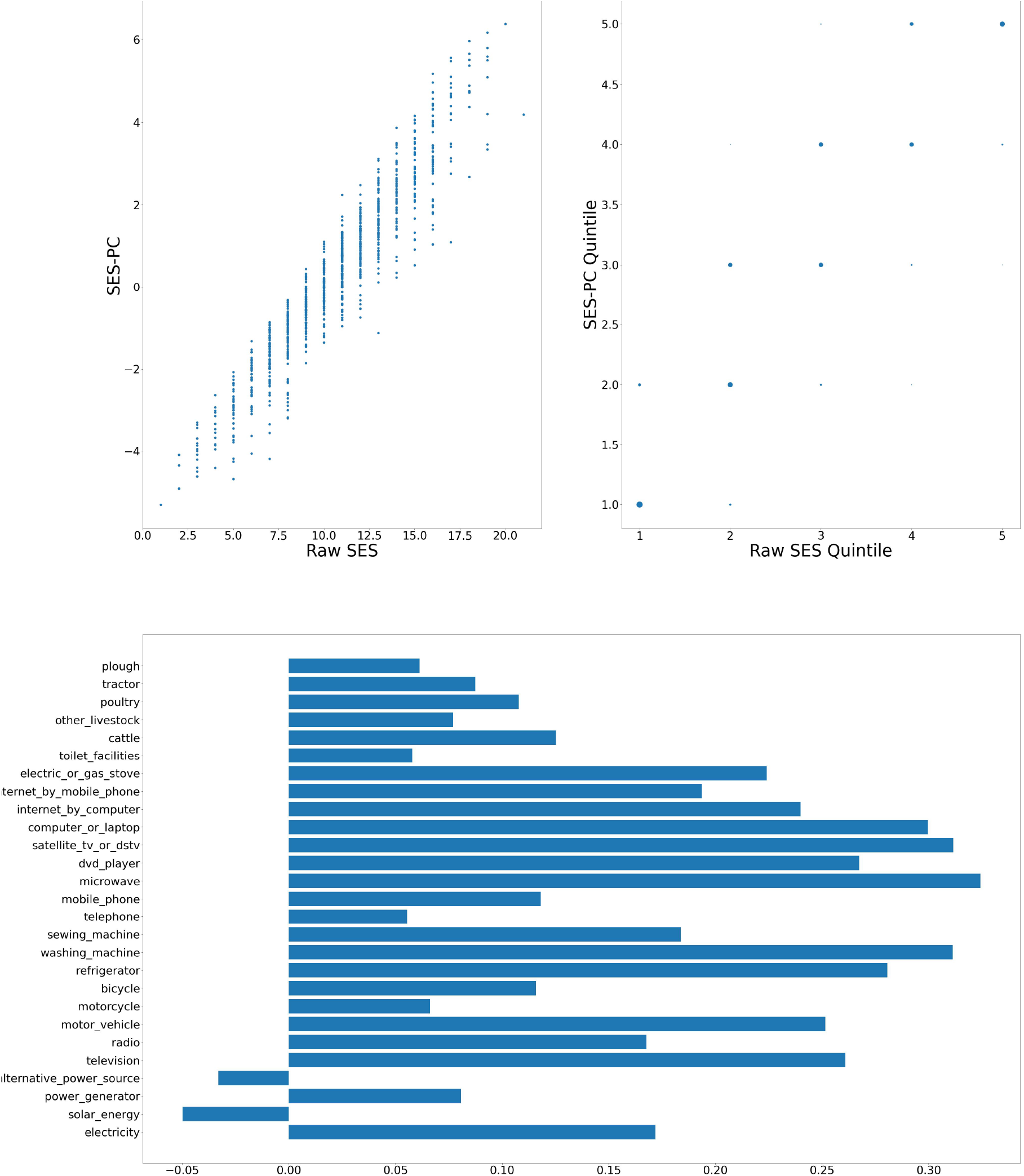
DIMAMO — Computation of SES score: graph at top left shows a scatter plot of the SES raw value versus adjusted value; top right shows the scatter plot of the computation quintiles using the different method with the size of the dot reflecting the number of individuals in each category; and at the bottom, the resulting weights of the assets for DIMAMO.

**Figure 3:**
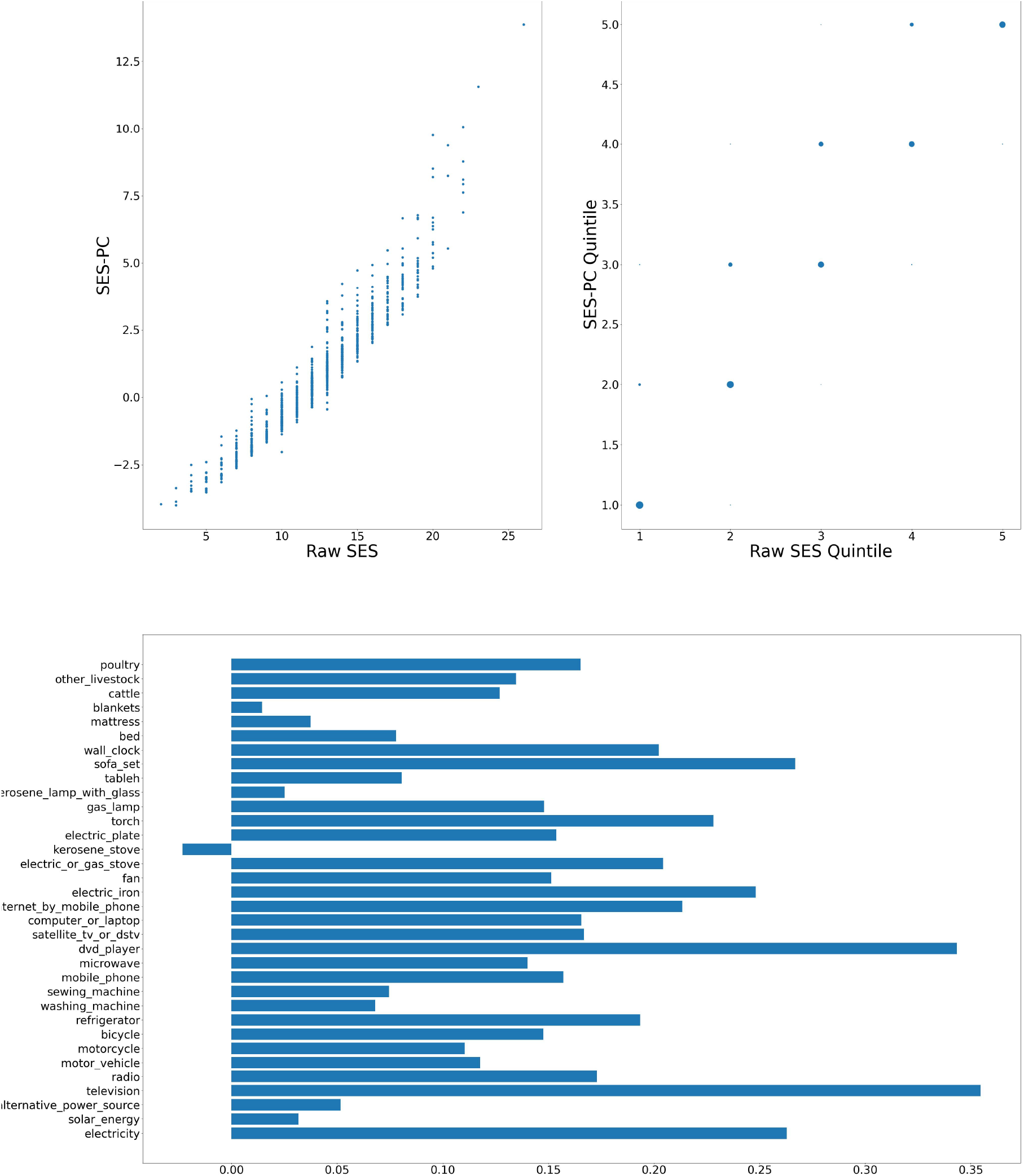
Nairobi — Computation of SES score: graph at top left shows a scatter plot of the SES raw value versus adjusted value; top right shows the scatter plot of the computation quintiles using the different method with the size of the dot reflecting the number of individuals in each category; and at the bottom, the resulting weights of the assets for Nairobi.

**Figure 4:**
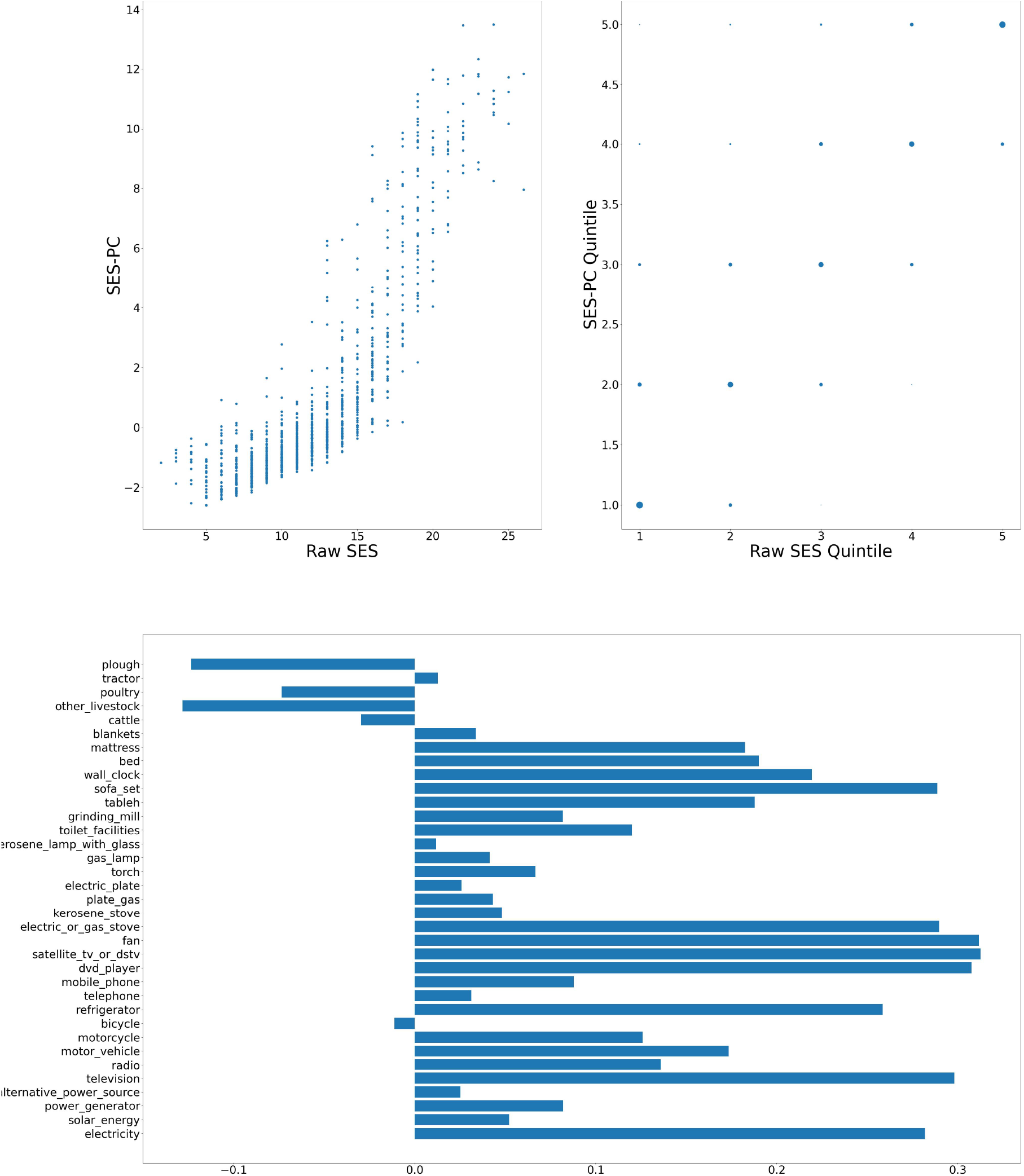
Nanoro — Computation of SES score: graph at top left shows a scatter plot of the SES raw value versus adjusted value; top right shows the scatter plot of the computation quintiles using the different method with the size of the dot reflecting the number of individuals in each category; and at the bottom, the resulting weights of the assets for Nanoro.

**Figure 5:**
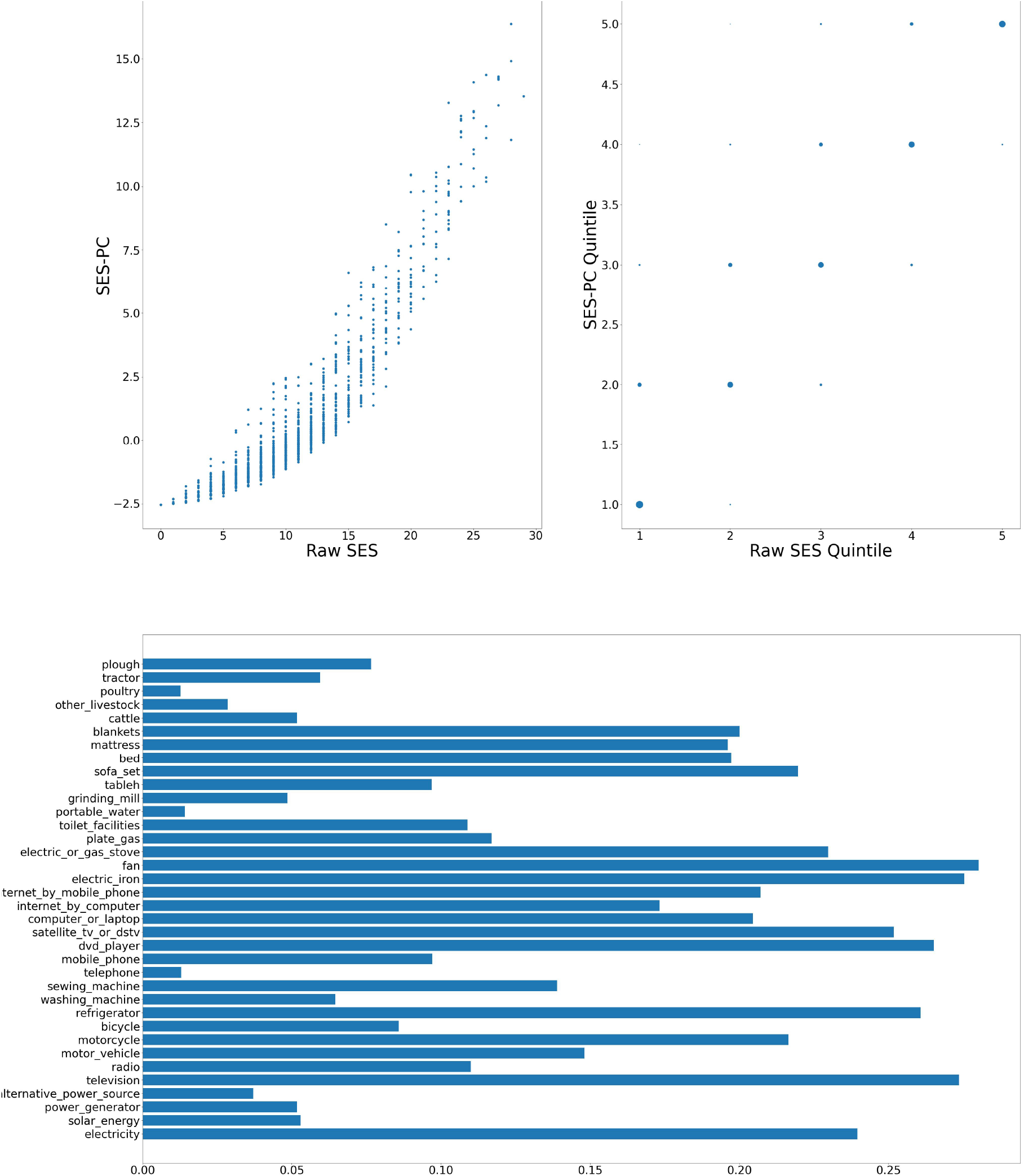
Navrongo — Computation of SES score: graph at top left shows a scatter plot of the SES raw value versus adjusted value; top right shows the scatter plot of the computation quintiles using the different method with the size of the dot reflecting the number of individuals in each category; and at the bottom, the resulting weights of the assets for Navrongo.

**Figure 6:**
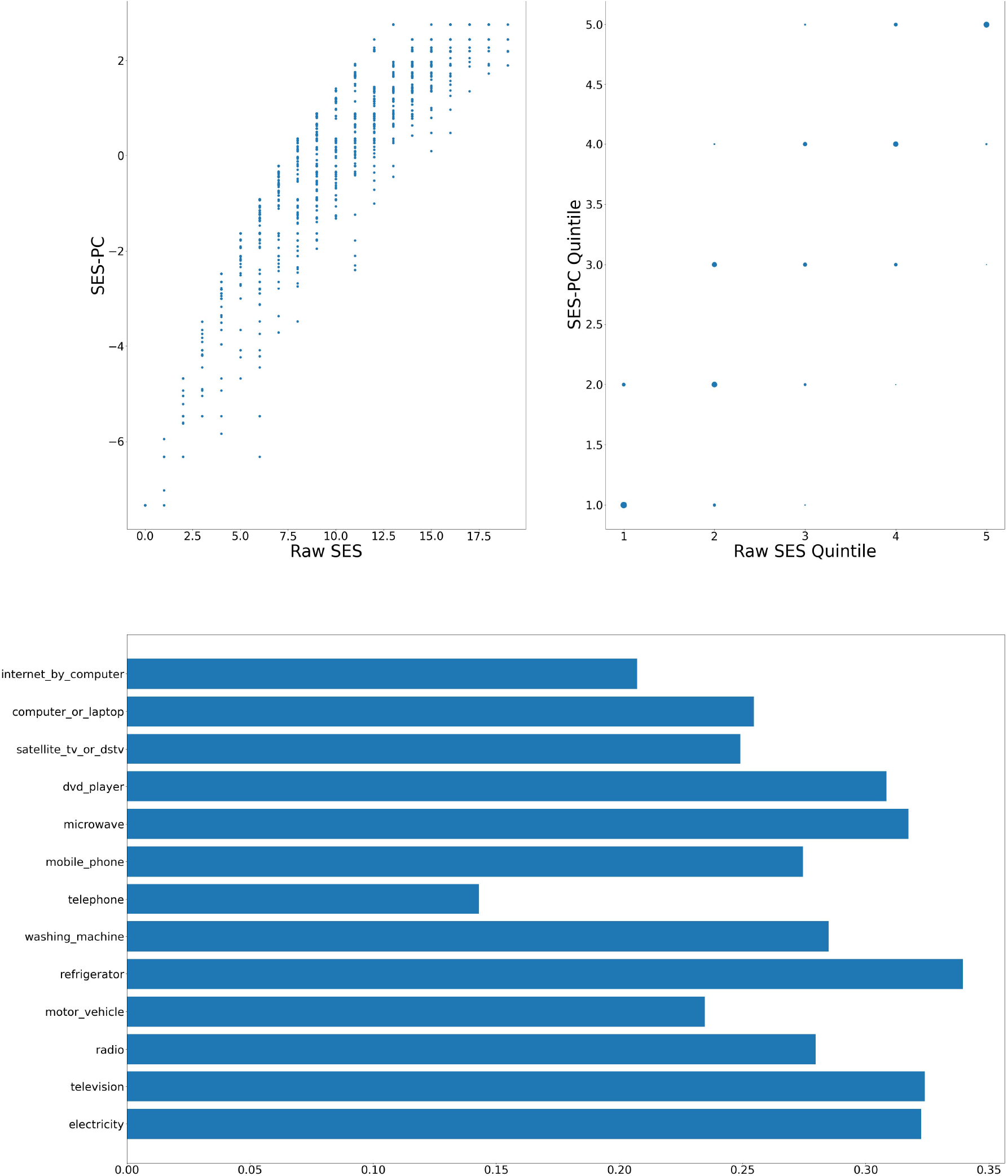
Soweto — Computation of SES score: graph at top left shows a scatter plot of the SES raw value versus adjusted value; top right shows the scatter plot of the computation quintiles using the different method with the size of the dot reflecting the number of individuals in each category; and at the bottom, the resulting weights of the assets for Soweto.

### 3.2. Principal component explanation of variance

Figure 7 shows the distribution of eigenvalues for the six AWI-Gen sites.

**Figure 7:**
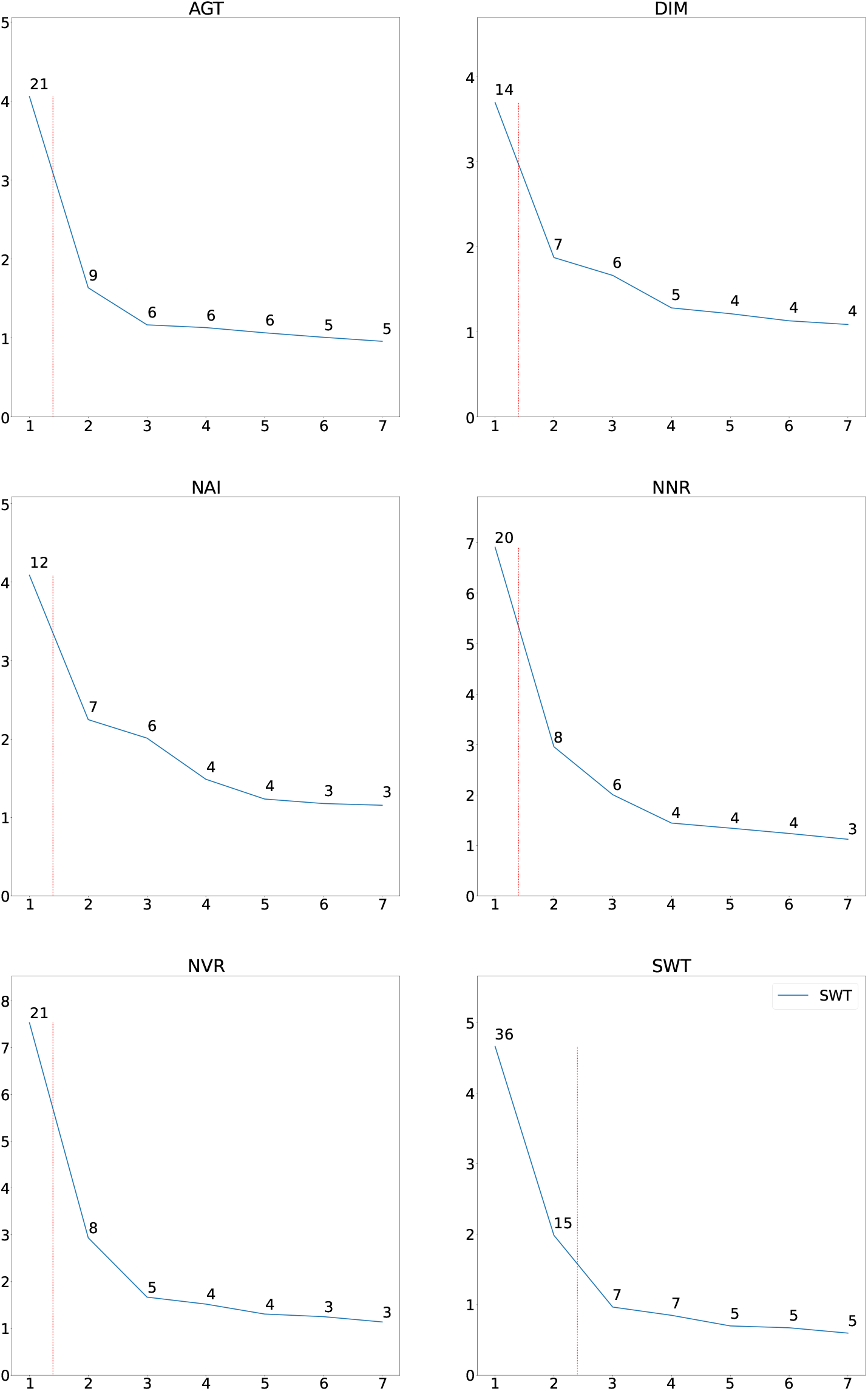
Distribution of eigenvalues for the six sites. The numbers of the eigenvectors are shown on the *x*-axis and the value of the corresponding eigenvalue is shown on the *y*-axis. The number annotating each point is the percentage variation that that eigenvalue explains. The red line shows the boundaries between the eigenvalues which explain at least 10% of the variation. Mostly only one PC explains at least 10%.

Only the first seven eigenvalues is shown. As can be seen a relatively small amount of the variation is explained by the first PC, except for Soweto.

## 4. Discussion and conclusion

We considered using the *pc*_*ses* due to its popularity. However, there were a number of factors that militated against it.

1. On strong theoretical grounds, Howe et al [6] and Kolenikov and Angeles [7] do not support this method for asset registers where the values are binary (present or absent) as they are in our case (and typically so in HDSS studies), without significant adjustment.
2. Hair et al [8] recommend that if this method is used, it is preferable to precede by consideration of whether some factors should be ignored based on their loadings, possible rotation of axes to reduce cross-loading, and the use of more than one principal component (it is common to use only the first principal component as in [2, 3] even though the PC explains only a small proportion of variance).
3. In particular, the PC method can and as was seen in Figure 4 does lead to negative values in the eigenvectors that are used in the factor analysis. For SES analysis, ownership of assets with negative eigenvectors reduces the socioeconomic status of the individual. In our case as an example, at the Nanoro site, ownership of a plough, poultry, livestock, or a bicycle would be negative (or alternatively and there is no theoretical way of distinguishing - these factors are positive and all others are negative). We cannot justify or support this conclusion, nor do we see it sensible to remove theseassets from the register.
4. The correlation between the raw SES scores and the PCA method of [2] when applied to AWI-Gen Phase I data was shown in Section 3.1. There is generally a high correlation between the scores. Nanoro, the site with the lowest correlation is characterised by having significant negative eigenvectors. Hence to the extent that the methods diverge, we prefer our approach, simplistic as it is.

For these reasons, we prefer the raw score to the PC-adjusted score.

## Data Availability

The AWI-Gen data set is available from the European Genome-phenome Archive (EGA) database (https://ega-archive.org/), with accession number EGAS00001002482 (phenotype dataset: EGAD00001006425). The availability of these datasets is subject to controlled access through, the Data and Biospecimen Access Committee of the H3Africa Consortium.

## Ethics

The Human Research (Medical) Ethics Committee of the University of the Witwatersrand, Johannesburg gave ethical approval for the AWI-Gen study (Protocol Numbers: M121029, M170880, M2210108). Each of the participating sites also obtained ethics approval from their respective ethics committees. AWI-Gen sample data was used as permitted by the informed consent provided by the study participants and according to the H3Africa policies and guidelines (www.h3africa.org).

